# Relation of incident Type 1 diabetes to recent COVID-19 infection: cohort study using e-health record linkage in Scotland

**DOI:** 10.1101/2022.02.11.22270785

**Authors:** Paul M McKeigue, Stuart McGurnaghan, Luke Blackbourn, Louise E Bath, David A McAllister, Thomas M Caparrotta, Sarah H Wild, Simon N Wood, Diane Stockton, Helen M Colhoun

## Abstract

**Background:** Studies using claims databases have reported that SARS-CoV-2 infection >30 days earlier increased the incidence of type 1 diabetes (T1DM). Using exact dates of type 1 diabetes diagnosis from the national register in Scotland linked to virology laboratory data we sought to replicate this finding.

**Methods:** A cohort of 1849411 individuals aged <35 years without diabetes, including all those in Scotland who subsequently tested positive for SARS-CoV-2, was followed from 1 March 2020 to 22 November 2021. Incident T1DM was identified by linkage to the national registry. Cox regression was used to test the association of time-updated infection with incident T1DM. Trends in incidence of T1DM in the total population from 2015-2021 were estimated in a generalized additive model.

**Findings:** There were 365080 in the cohort with at least one detected SARS-CoV-2 infection during follow-up and 1074 who developed T1DM. The rate ratio for incident T1DM associated with first positive test for SARS-CoV-2 (with no previous infection as reference category) was 0.88 (95% CI 0.63 to 1.23) for infection more than 30 days earlier and 2.62 (95% CI 1.81 to 3.79) for infection in the previous 30 days. However negative and positive SARS-CoV-2 tests were more frequent in the days surrounding T1DM presentation. In those aged 0-14 years incidence of T1DM during 2020-2021 was 20% higher than the 7-year average.

**Interpretation:** T1DM incidence in children increased during the pandemic. However the cohort analysis does not support a causal effect of SARS-CoV-2 infection itself on T1DM incidence.

**Funding:** None

## Background

A recent paper from the US Centers for Disease Control and Prevention (CDC) based on medical claims databases [1] reported increased incidence of diabetes 30 days or more after SARS-CoV-2 infection among those aged under 18 years. Such a finding, if replicated, would imply a substantial increase in the burden of childhood onset diabetes and might also alter the risk/benefit balance for COVID-19 vaccination in young children. The CDC study did not distinguish the type of diabetes but in this age group type 1 diabetes accounts for about 75% of cases in the US [2]. There is evidence that viral infections can play a primary causal role in type 1 diabetes through infection of pancreatic beta-cells, triggering auto-immunity [3] and that viral infections can have a secondary causal role through accelerated auto-immunity increasing the rate of progression to stage 3 type 1 diabetes in those with autoimmune beta-cell destruction who are still normoglycaemic. However there are other possible explanations for the observed association:

- misclassification of prevalent diabetes as incident in people who acquire SARS-CoV-2 infection; this is plausible given that type 1 diabetes is already known to be associated with increased risk of COVID-19 [4].
- COVID-19 may cause diabetes to be diagnosed earlier, through precipitating metabolic decompensation or simply through causing urine or blood to be tested for diabetes.
- early symptoms of diabetes may lead to contact with health services and to testing for SARS-CoV-2, and thus to detection of infections that would otherwise go undetected.

In Scotland health care is free at the point of delivery and all new diagnoses of type 1 diabetes in primary or secondary care are captured into the nationwide diabetes Scottish Care Information-Diabetes (SCI-Diabetes) registry [5] within 24 hours of the diagnostic care encounter. Furthermore the nationwide policy is that all paediatric (<16 years) cases of suspected diabetes must be admitted for inpatient care to a specialist unit on the day of presentation. Adults with suspected type 1 diabetes are referred urgently to a hospital diabetes clinics and are seen within a few days. All PCR tests for COVID-19 have been captured into a national database by Public Health Scotland. This makes it possible to determine the temporal sequence of the relationship between SARS-CoV-2 infection and diagnosis of type 1 diabetes based on exact dates of first positive test and accurate dates of diagnosis of type 1 diabetes. The objectives of this study were:

- to establish whether SARS-CoV-2 infection is associated with increased risk of incident Type 1 diabetes
- to determine the time interval over which risk is increased
- to examine how the incidence of type 1 diabetes has changed during the pandemic.

## Methods

The REACT-SCOT study, described in detail elsewhere [6,7], was established by Public Health Scotland early in the epidemic, as a matched case-control study of all diagnosed COVID-19 cases in Scotland with up to ten controls per case, matched for age, sex and general practice. The incidence density sampling design allows individuals to be sampled more than once as a control and subsequently as a case. The 3938454 individuals sampled in the REACT-SCOT study up to 22 November 2021 comprise 72% of the estimated Scottish population in mid-2020, including all diagnosed cases of COVID-19. The cohort at risk of diabetes was formed from all 1849411 individuals sampled in REACT-SCOT who at baseline were aged less than 35 years and had not been diagnosed with diabetes: Supplementary Figure S1 shows a flow chart of the construction of the cohort. The baseline (entry) date for each individual was the later of 1 March 2020 (the start of the epidemic in Scotland) or the date of birth. Date of exit was the earliest of: latest date of extraction of linked data (22 November 2021), date of death, or date of any evidence of diabetes (dispensed prescription for a drug used in diabetes, hospital admission with discharge diagnoses including ICD-10 code E10 to E14, or an outpatient consultation with specialty coded A81 for diabetes. COVID-19 vaccination records were obtained from a national database (the National Clinical Data Store).

### Ascertainment of SARS-CoV-2 infection

SARS-CoV-2 infection was defined by first positive PCR test for SARS-CoV-2 in the Electronic Communication of Surveillance in Scotland (ECOSS) database that captures all PCR tests for COVID-19 nationally. REACT-SCOT cases include some individuals with a definite clinical diagnosis of COVID-19 (ascertained via hospital discharge diagnosis coding) who had never tested positive. These test-negative cases amounted to only 1% of COVID-19 cases in the cohort at risk of diabetes: date of presentation with SARS-CoV-2 infection for these cases was set as 7 days before admission. Suspected reinfection was defined by the CDC criterion of a positive test at least 90 days after the first positive test [8].

### Diabetes ascertainment

Incident cases of type 1 diabetes in Scotland during 2015-2021 were ascertained based on date of diagnosis and type of diabetes as recorded by the clinician in the Scottish Care Information-Diabetes (SCI-Diabetes) registry. The clinical classification of type in SCI-Diabetes has previously been validated against detailed prescribing and hospital admission histories; in the years 2015-2019 we reclassified the type of diabetes as type 2, monogenic or secondary in less than 2% of those aged under 16 clinically labelled as type 1 diabetes for example.

### Statistical analysis

Incident cases of type 1 diabetes during the study period 1 March 2020 to 22 November 2021 were linked to the REACT-SCOT database. Those with pre-existing diabetes of any type at baseline were excluded using SCI-Diabetes records, supplemented with prescribing data (240-day lookback for British National Formulary subparagraph codes 0601011 or 0601012 for drugs for diabetes), hospital discharge diagnoses (5-year lookback for any mention of ICD-10 codes E10 to E14 for diabetes) and outpatient records (any consultation with specialty code A81 for diabetes). As shown in Table S1 almost all incident cases have a first admission for diabetes within a few days of the diabetes diagnosis date.

The association of Type 1 diabetes with recent SARS-CoV-2 in REACT-SCOT was modelled in a Cox regression. COVID-19 status at the start of each person-time interval was coded as no prior COVID-19, first positive test in the last 30 days, or first positive test >30 days earlier. Age at baseline was modelled as a natural spline with three degrees of freedom. Other covariates included in the model were sex and number of vaccine doses (0 to 3) given at least 14 days before. There were no missing values. To allow exact updating of time-varying covariates – SARS-CoV-2 status and vaccination status – observed person-time was split into 1-day intervals. Without thinning, this time-splitting would generate a data table with more than 1 billion rows. To reduce computational requirements, the rows were thinned after splitting, to retain a 1% sample of person-time intervals with no event and all person-time intervals with an event. There is no appreciable loss of information from this procedure as even after thinning there are still more than 10,000 controls (person-time intervals with no event) for every case (person-time intervals with an event). Ascertainment bias was explored by examining patterns of both negative and positive tests in relation to diagnosis date.

A table of incident cases of type 1 diabetes in Scotland from 2015 to end of 2021 was joined by date and age group to daily estimates of the total population of Scotland, obtained by fitting a spline function to publicly-available mid-year estimates. Incidence in a 56-day sliding window centred on each day from 1 March 2015 to 1 October 2021 was calculated for age bands 0-14 and 15-34 years. Smoothed effects of seasonality and calendar time were estimated jointly from the counts of daily cases by age group using the R package mgcv [9] to fit a generalized additive model as detailed in the Supplementary Appendix. PmcK conducted the data analysis.

### Ethics approval and data governance

Approval for use of the diabetes data was provided by the Public Benefit and Privacy Panel (https://www.informationgovernance.scot.nhs.uk/pbpphsc/) (ref. 1617-0147) and the Scotland A Multicentre Research Ethics Committee (ref 21/WS/0047). The REACT-SCOT study was performed within Public Health Scotland as part of its statutory duty to monitor and investigate public health problems and pandemic response. Under the [UK Policy Framework for Health and Social Care Research](https://www.hra.nhs.uk/planning-and-improving-research/policies-standards-legislation/uk-policy-framework-health-social-care-research/) set out by the NHS Health Research Authority, this does not fall within the definition of research and ethical review was therefore not required. This has been confirmed in writing by the NHS West of Scotland Research Ethics Service. Individual consent is not required for Public Health Scotland staff to process personal data to perform specific tasks in the public interest that fall within its statutory role. The statutory basis for this is set out in Public Health Scotland’s [privacy notice](https://www.publichealthscotland.scot/our-privacy-notice/personal-data-processing/).

### Funding

No specific funding was received for this study. HC is supported by an endowed chair from the AXA Research Foundation. TM is supported by Diabetes UK grant 18/0005786. Authors were not precluded from accessing data in the study, and they accept responsibility to submit for publication.

## Results

### Association of SARS-CoV-2 infection with incident type 1 diabetes

Among the 1849411 individuals in the cohort there were 365080 with a first detected SARS-CoV-2 infection between 1 March 2020 and 22 November 2021. There were 1074 persons who developed incident type 1 diabetes during follow up. 1052 persons were right censored for having developed other types of diabetes and 447 due to death before the end of the study period. Altogether there were 3.16 million persons years in the analysis (3.01 million unexposed to SARS-CoV-2).

The age distribution of persons in the cohort who developed type 1 diabetes and their COVID-19 status on or prior to their date of diagnosis of diabetes is shown in Table 1. Overall SARS-CoV-2 infection had been detected in 18 (2%) of incident cases of type 1 diabetes up to their date of diagnosis. Just two of these tested positive again at least 90 days after their first positive test, meeting CDC criteria for suspected reinfection [8].

**Table 1.** SARS-CoV-2 infection status at or before diagnosis in those diagnosed with type 1 diabetes during follow-up, by age at entry to cohort.

| Age at entry | SARS-CoV-2 status |  |  |  |  |
| --- | --- | --- | --- | --- | --- |
|  | No positive test | 0 to 15 days | 16 to 30 days | >30 days | All |
| 0 to <16 | 592 (93%) | 14 (2%) | 10 (2%) | 19 (3%) | 635 |
| 16 to <35 | 413 (94%) | 4 (1%) | 3 (1%) | 19 (4%) | 439 |
| All | 1005 (94%) | 18 (2%) | 13 (1%) | 38 (4%) | 1074 |

After splitting into 1-day person-time intervals and thinning as described above, the dataset used for Cox regression consisted of 11553301 person-time intervals. Table 2 shows the hazard ratio for incident type 1 diabetes by COVID-19 status categorized as to timing of the SARS-CoV-2 infection. With no previous infection as reference category, the hazard ratios for type 1 diabetes associated with prior SARS-CoV-2 infection were 2.62 (95% CI 1.81 to 3.79) for infection in the last 30 days, and 0.88 (95% CI 0.63 to 1.23) for first positive test more than 30 days earlier. In those aged less than 16 years the corresponding rate ratios were 3.15 (95% CI 2.07 to 4.79) and 0.81 (95% CI 0.51 to 1.30). COVID-19 vaccination status was not associated with incidence of type 1 diabetes.

**Table 2.** Rate ratios for new diagnosis of Type 1 diabetes associated with time since testing positive for SARS-CoV-2.

| Effect | No diabetes<br>(11552227) | Incident type 1 (1074) | Rate ratio<br>(95% CI) | *p*-value |
| --- | --- | --- | --- | --- |
| *Male sex* | 5806341 (50%) | 617 (57%) | 1.33 (1.17, 1.50) | $5 \times 10^{-6}$ |
| $\geq 1$ dose vaccine | 1376686 (12%) | 106 (10%) | 0.93 (0.73, 1.18) | 0.5 |
| *Days since first testing positive for SARS-CoV-2* |  |  |  |  |
| No positive test | 11007838 (95%) | 1005 (94%) | . | . |
| 0 to 30 days | 106460 (1%) | 31 (3%) | 2.62 (1.81, 3.79) | $3 \times 10^{-7}$ |
| >30 days | 437929 (4%) | 38 (4%) | 0.88 (0.63, 1.23) | 0.4 |

Cox regression model with person-time split into 1-day intervals
Person-time intervals with no event thinned to a 1% random sample to reduce computation
Vaccination and SARS-CoV-2 status coded as time-updated covariates
Age at entry included in the model as a natural spline with 3 degrees of freedom

### Pattern of testing for COVID-19 in relation to type 1 diabetes presentation

To investigate if the association of incident diabetes with a first positive test in the last 30 days could be explained by higher testing rates around the date of diagnosis of diabetes, we plotted the dates of all PCR tests (negative or positive), in people with incident type 1 diabetes. As shown in Figures S2 and 1 there was marked increased frequency of testing, mostly negative, in the days before and after diagnosis of type 1 diabetes.

### Population level incidence rates of type 1 diabetes

Figure 2 shows the observed incidence of type 1 diabetes in the Scottish population aged less than 35 years over 56-day sliding time windows during 2015-2021. The age bands 0-14 and 15-34 were used for compatibility with the age bands used for national population estimates. A pattern of peaks and troughs is seen in both age groups having greater amplitude in the 0-14 age band. To discern underlying patterns requires modelling of seasonality and calendar time effects as described in the Supplementary Appendix. Supplementary Figure S3 (a) shows the fitted seasonal effect as the ratio of incidence per week of the year to the average rate, adjusted for calendar time. Incidence peaks in February and September, and is lowest in July. Supplementary Figure S3 (b) shows the fitted effect of calendar time from 2015-2021 as the ratio of the incidence per day to the average rate, adjusted for seasonality. In those aged 15-34 years the relationship best supported by the data is a linear increase over 2015-2021. In those aged 0-14 years the relationship best supported by the data is a curve with considerable year to year variation, with a trough in mid-2019 at 0.9 times the long-term average followed by a rise to a peak in 2021 of 1.2 times the long-term average.

**Fig 1.**
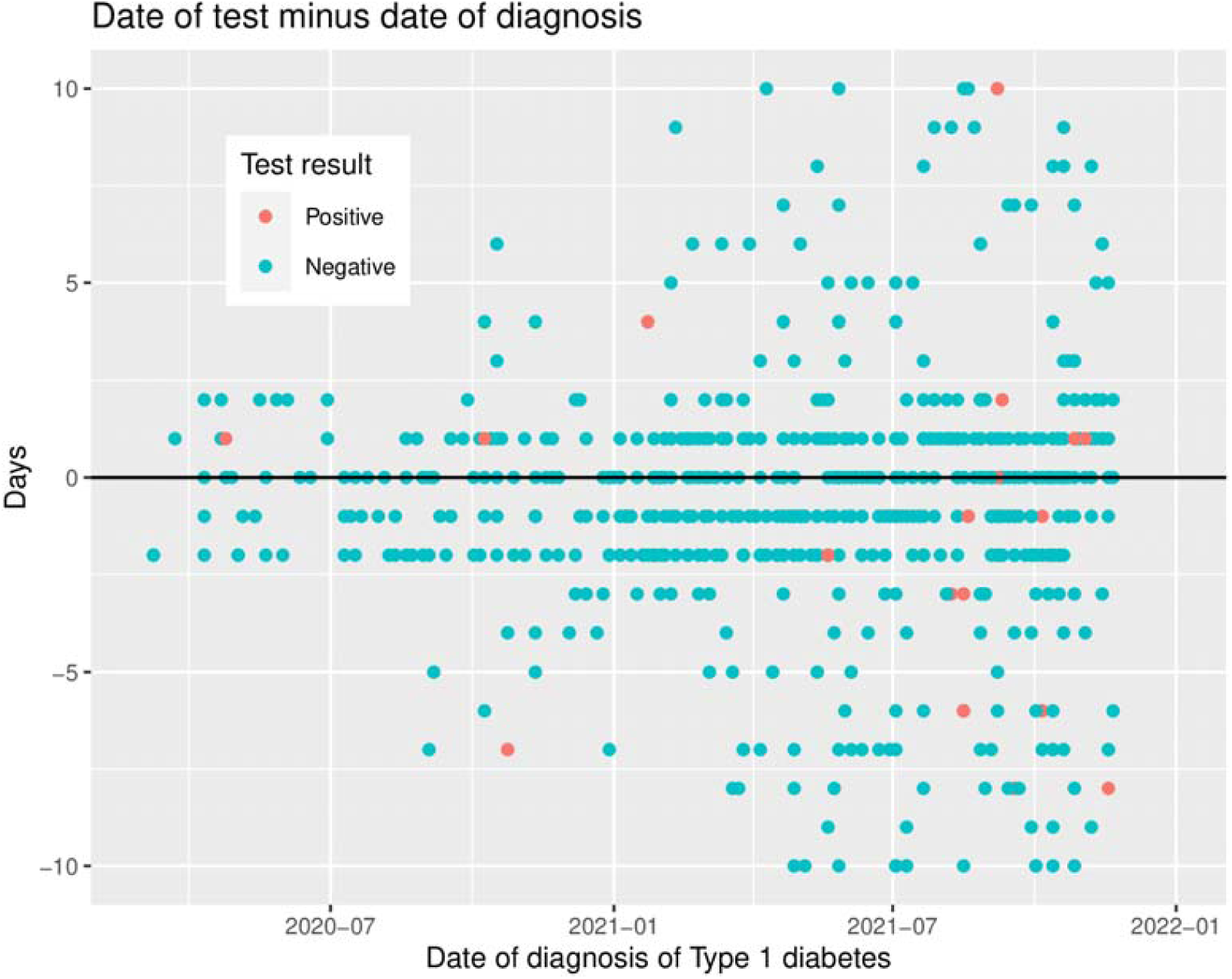
Dates of SARS-CoV-2 PCR tests minus date of diagnosis: zoomed plot from 10 days before to 10 days after date of diagnosis.

**Fig 2.**
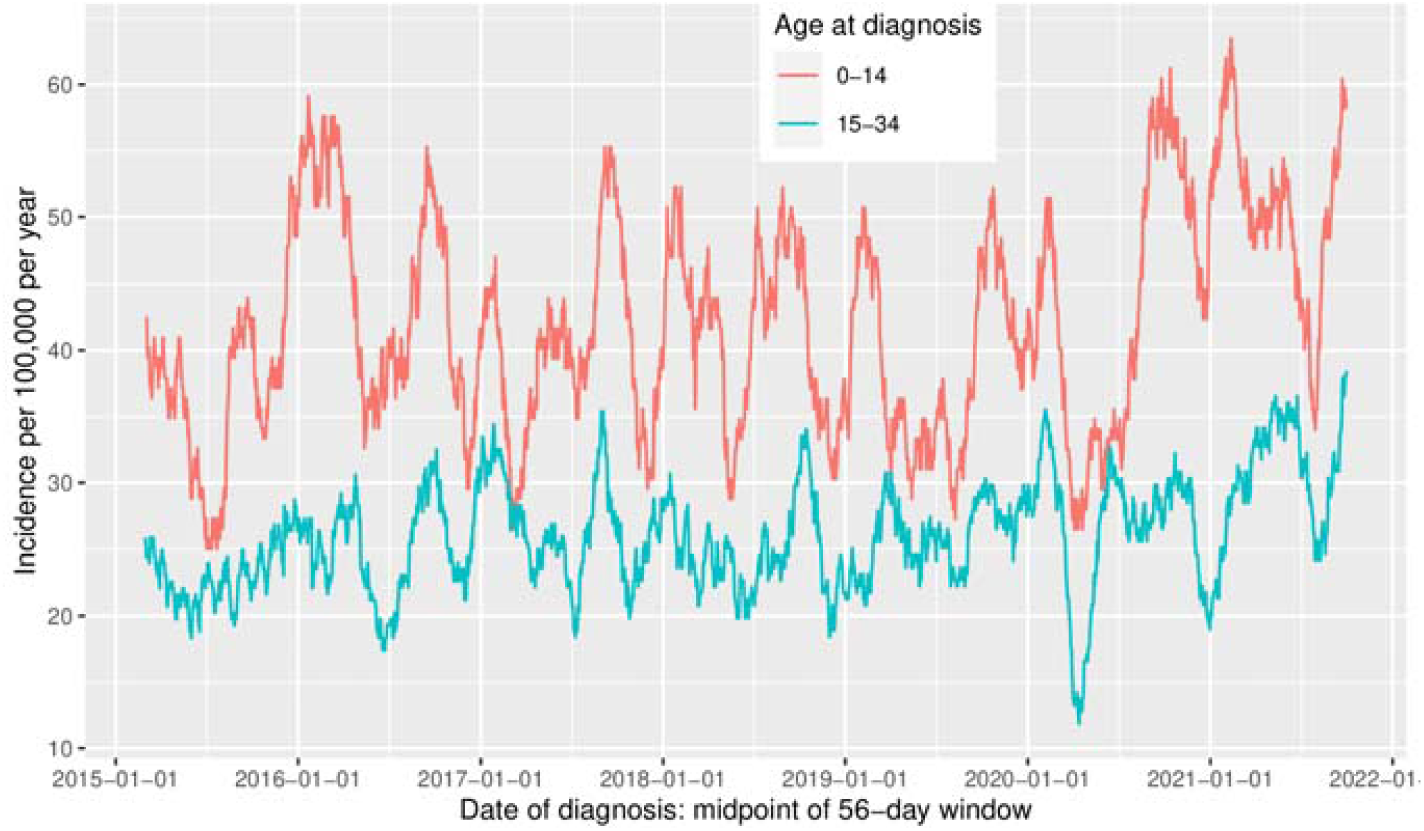
Incidence of type 1 diabetes in Scotland: sliding time windows of width 56 days, by age at diagnosis.

## Discussion

### Statement of principal findings

In this study using precise dates of diagnosis for type 1 diabetes we found that there was no association of SARS-CoV-2 infection >30 days previously with type 1 diabetes incidence overall or in those aged <16 years. We observed a strong association between a first positive SARS-CoV-2 tests within the past 30 days and type 1 diabetes in both children and adults. Some of this association may be attributable to COVID-19 illness causing metabolic decompensation and precipitating diagnosis of incipient type 1 diabetes. In England the median time from onset of symptoms to diagnosis of type 1 diabetes before age 16 years was reported to be 25 days [10]. As the median incubation period of SARS-CoV-2 is less than 5 days, it is likely that symptoms of diabetes preceded the infection in most of those who first tested positive for SARS-CoV-2 less than 20 days before diagnosis of type 1 diabetes. However, the increased frequency of recent negative as well as positive SARS-CoV-2 tests around the time of presentation with type 1 diabetes indicates that the association is partly attributable to higher detection of infection through increased testing around the time of presentation with type 1 diabetes. The absence of any protection from COVID-19 vaccination against type 1 diabetes is further evidence against a causal effect of COVID-19 on type 1 diabetes.

The estimated incidence of type 1 diabetes in those aged 0-14 years in Scotland increased during 2020-2021 to about 1.2 times the average over the study period. However in this age group incidence varies widely from year to year with 2019 for example having 0.9 times the long term average incidence. In those aged 15-34 years there was no evidence of an increase in 2020-2021 beyond the long term upward trend in this age band. Increased incidence of type 1 diabetes could have several possible explanations other than SARS-CoV-2 infection itself. Of note the levels of infection with other respiratory viruses has been altered over this period [11,12] and it is plausible that there may be changes in other pathogens such as enteroviruses that have been associated with altered risk of type 1 diabetes [13–17]. Changes in other environmental factors such as sunlight exposure and vitamin D levels might also be relevant. [18]. It should be noted that the seasonal pattern with a peak in incidence in February and September is the same as previously reported for Denmark [19] and here we demonstrate it is observed in older onset as well as childhood onset type 1 diabetes though at lower amplitude. It is of interest that the seasonal pattern was maintained during the pandemic despite altered seasonal social mixing patterns.

### Strengths and limitations

Strengths of this study include the availability of individual level data, comprehensive national coverage of PCR tests, the inclusion of data on the level of negative testing around the time of presentation and most importantly validation of the accuracy of dates of diagnosis of type 1 diabetes in the SCI-diabetes registry against date of first hospitalization for type 1 diabetes in paediatric cases for whom the policy is to admit all newly-diagnosed cases immediately. Limitations are that because the numbers of incident cases of type 1 diabetes exposed to SARS-CoV-2 infection were relatively small, for formal modelling of the hazard ratio we had to use fairly broad categories of 0-30 and >30 days for exposure period. However the clustering of negative and positive tests around the date of diagnosis is obvious on inspection of scatter plots. Another limitation common to other studies of this question are that until mass testing was rolled out in late 2020, most cases of SARS-CoV-2 in younger people were not detected. However the cumulative incidence of infection in 5-14 year olds in the UK is estimated to have been only about 15% up to late 2020 [20], so misclassification of exposed individuals as unexposed would only slightly reduce the rate ratios for Type 1 diabetes associated with detected infection in this age group.

### Comparison with previous studies

Our results do not confirm the association of incident diabetes before age 18 years with SARS-CoV-2 infection more than 30 days previously, reported by CDC [1]. In that study the rate ratio associated with COVID-19 exposure was estimated as 2.7 in the IQVIA database and 1.3 in the Health Verity database. It is not clear how accurately the date of diagnosis could be determined from these claims databases. The incidence rates of 337 in the IQVIA dataset and 351 per 100,000 in the HealthVerity dataset are far higher than the most recent incidence rates reported from the SEARCH registry in the US: 22 per 100,000 for type 1 diabetes in those aged 0-19 years and 5 per 100,000 for type 2 diabetes in those aged 10-19 years [21], and far higher than the rates in Scotland. Some cases classified as incident in the CDC study on the basis of a first encounter for diabetes had a prior history of diabetic ketoacidosis. These aspects suggest that some prevalent cases may have been misclassified as incident. No other studies to our knowledge have directly tested the association at individual level of SARS-CoV-2 infection with type 1 diabetes onset.

Two studies have reported increased incidence of type 1 diabetes during the pandemic. A cohort study in the Finnish Paediatric Diabetes Registry reported that annual incidence had increased from an average of 39 in the 2016-2019 period to 56 per 100,000 during April-October 2020 [22]. Of the 33/84 new cases during the pandemic who were tested for SARS-CoV-2 IgG, all were negative suggesting a cause other than COVID-19 itself A Romanian registry also reported a temporal increase in incident T1D during the pandemic period of 13.3/100,000 compared to 11.0-12.3/100,000 for the 2015-2019 period. [23]. In contrast, studies from Germany [24], Saudi Arabia [25] and Italy [26] have found incidence to be no higher during the pandemic than before. It would be of interest to further establish incidence changes in other countries and to map changes in incidence by country against SARS-COV-2 infection rates and the stringency of social distancing measures.

## Conclusion

In conclusion, we observed during the pandemic a 1.2 fold increase in the incidence of type 1 diabetes in those aged 0-14 years; there are several possible causes for this. In a cohort analysis, incident type 1 diabetes was not associated with SARS-CoV-2 infection more than 30 days previously so that a causal effect of SARS-CoV-2 is not supported.

## Data Availability

The component datasets used here are available to researchers via application to the Public Benefits and Privacy Panel for Health and Social Care https://www.informationgovernance.scot.nhs.uk/pbpphsc/

## Declarations

### Public and Patient Involvement statement

This study was conducted under approvals from the Public Benefit and Privacy Panel for Health and SocialCare which includes public and patient representatives.

### Data Availability

The component datasets used here are available to researchers via application to the Public Benefits and Privacy Panel for Health and Social Care (https://www.informationgovernance.scot.nhs.uk/pbpphsc/). All final source code used for derivation of variables, statistical analysis and generation of this manuscript can be accessed at https://github.com/pmckeigue/covid-scotland_public.

### Contribution Statement

PM and HC conceived and designed the study and made the decision to submit. HC, SM, LB, DM and SW contributed to retrieving or pre-processing data for the study. TC undertook literature searches. LB contributed to data interpretation. PM accessed and analysed the data. SNW oversaw the time-series modelling. HC accessed and verified the analysis and wrote the first draft of the manuscript. All authors contributed to reviewing and editing the manuscript for intellectual content. All authors approved the decision to submit.

