## Supplementary appendix for "Relation of incident Type 1 diabetes to recent COVID-19 infection: cohort study using e-health record linkage in Scotland"

#### Validation of date of diagnosis against date of admission

As policy in Scotland is to admit immediately anyone with a suspected diagnosis of type 1 diabetes before age 16 years, the date of admission recorded in Scottish Morbidity Record (SMR) 01 can be used to validate the accuracy of date of diagnosis recorded in SCI-Diabetes. Table S1 tabulates the difference between date of diagnosis of type 1 diabetes recorded in SCI-Diabetes and the date of first admission for diabetes recorded in SMR01. In those aged less than 16 years, 86% (348 of 403) of dates of diagnosis recorded in SCI-Diabetes were within two days of the date of first admission for diabetes. For cases with date of encashed insulin prescription, date of admission for diabetes or date of outpatient consultation earlier than the date of diagnosis recorded in SCI-Diabetes, the date of diagnosis was recoded as the earliest date of these dates.

#### Generalized additive model of type 1 diabetes incidence during 2015-2021

The R package mgcv [1] was used to fit a generalized additive model with Poisson likelihood to the counts of daily cases by age band from 2015 to 2021. The terms in the model were the intercept, an indicator for weekend versus weekday, a smoothed term for seasonality as a cyclic cubic spline fitted to the week of diagnosis (encoded as an integer from 1 to 52), and a smoothed term for calendar time as a thin plate spline fitted to the date of diagnosis. These smoothed terms were specified separately for each of the two age bands, and are defined to have zero mean within each age band. The model-fitting routine chooses the effective number of degrees of freedom for each spline by minimizing the unbiased risk estimator (penalized deviance). In those aged 0-14 years at diagnosis, the effective number of degrees of freedom was 9.6 for the seasonality effect and 10 for the calendar time effect. In those aged 15-34 years, the effective number of degrees of freedom was 4.5 for seasonality and 1.2 for calendar time (nearly equivalent to a linear effect). Figure S3 shows the fitted curves for seasonality (panel a) and calendar time (panel b) by age band. Though the seasonality effect in those aged 15-34 is not statistically significant when tested in isolation, the peaks and troughs of the curve estimated in this age group coincide with those of the curve fitted to the 0-14 age group.

### Fig S1. Flow chart of construction of cohort at risk of incident diabetes from REACT-SCOT sample

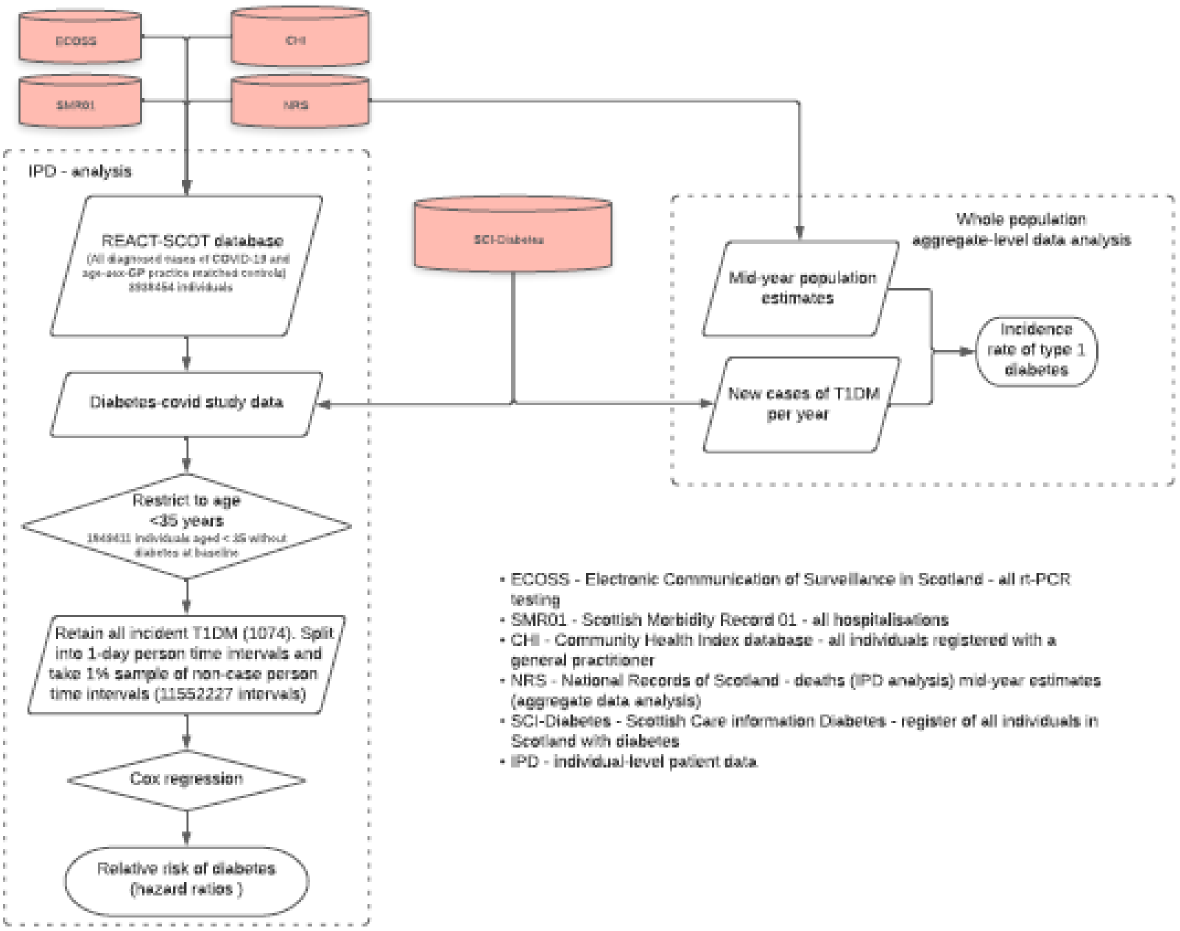

#### Fig S2. Relation of date of SARS-CoV-2 tests to date of diagnosis of type 1 diabetes.

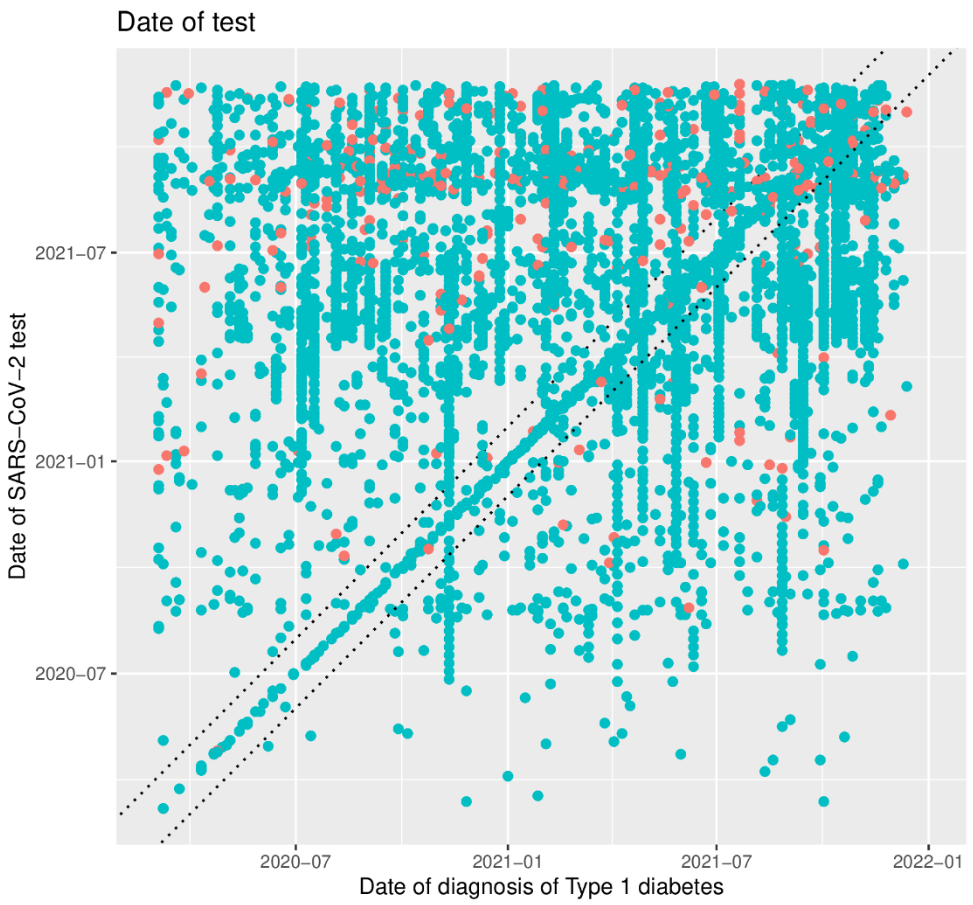

The dotted lines bound the interval from 10 days before to 10 days after diagnosis

#### Fig S3. Fitted curves for relation of type 1 diabetes incidence in Scotland to seasonality and calendar time from 2015 to 2021.

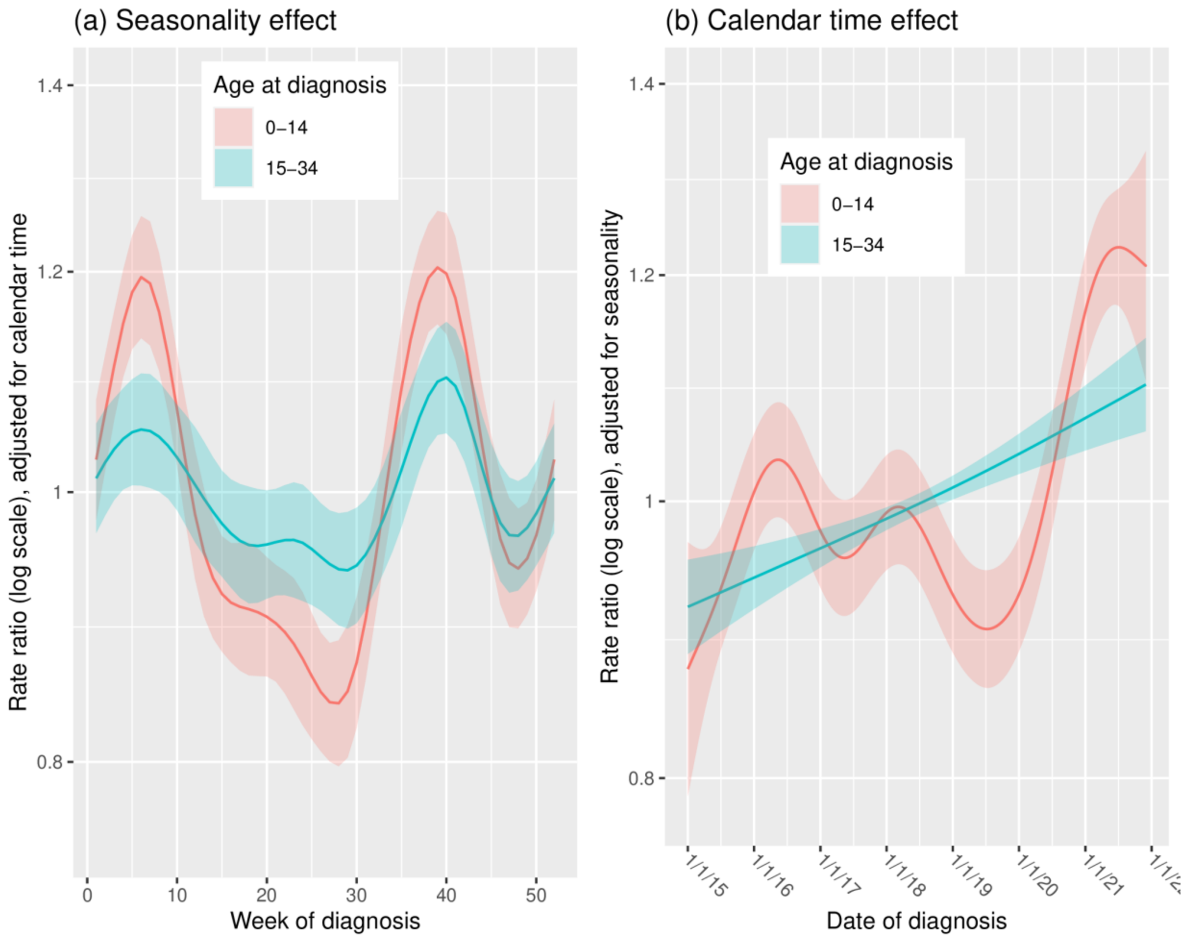

Ribbon edges are one standard error above and below the fitted curve

#### **Table S1.** Days from first admission for diabetes to date of diagnosis of Type 1 diabetes recorded in SCI-Diabetes, by age at entry to cohort

| Date of diagnosis in SCI-Diabetes | Age at entry (years) | |
|---|---|---|
| | <16 | 16 to <35 |
| 3 or more days before admission | 36 (9%) | 23 (11%) |
| Within 2 days | 368 (88%) | 161 (79%) |
| 3 or more days after admission | 12 (3%) | 19 (9%) |
| Total | 416 | 203 |

SMR01 hospital admission records were not available for the most recently-diagnosed cases, as these records are updated only once every 1-2 months
